# Utilizing Large Language Models to Simplify Radiology Reports: a comparative analysis of ChatGPT3.5, ChatGPT4.0, Google Bard, and Microsoft Bing

**DOI:** 10.1101/2023.06.04.23290786

**Authors:** Rushabh Doshi, Kanhai Amin, Pavan Khosla, Simar Bajaj, Sophie Chheang, Howard P. Forman

## Abstract

This paper investigates the application of Large Language Models (LLMs), specifically OpenAI’s ChatGPT3.5, ChatGPT4.0, Google Bard, and Microsoft Bing, in simplifying radiology reports, thus potentially enhancing patient understanding. We examined 254 anonymized radiology reports from diverse examination types and used three different prompts to guide the LLMs’ simplification processes. The resulting simplified reports were evaluated using four established readability indices. All LLMs significantly simplified the reports, but performance varied based on the prompt used and the specific model. The ChatGPT models performed best when additional context was provided (i.e., specifying user as a patient or requesting simplification at the 7th grade level). Our findings suggest that LLMs can effectively simplify radiology reports, although improvements are needed to ensure accurate clinical representation and optimal readability. These models have the potential to improve patient health literacy, patient-provider communication, and ultimately, health outcomes.

## Introduction

Imaging reports are a cornerstone of medical decision-making, providing information for diagnosis, treatment planning, and monitoring disease progression. Historically, only the radiologist and referring provider accessed these reports, but the rise of telemedicine and patient portals, as well as regulatory changes, most recently the 21st Century Cures Act, have increased access to electronic health records and transformed patients’ relationship with their medical information.^1–4^

Digital health literacy, defined as the degree to which a patient can obtain, process, and understand electronic information,^5^ is critical for patients to fully benefit from this transformation.^6^ Radiology reports, however, are filled with technical jargon, making them relatively uninterpretable to individuals without a clinical background.^7^ Expanded access to these reports could thus exacerbate patient anxiety, misunderstanding, and emotional distress, particularly with abnormal findings.^8–10^ Improving radiological literacy could help address these concerns, with other spillover benefits to safety and transparency,^11^ shared decision-making,^12^ treatment compliance,^13^ and reducing health disparities.^14^

Fifteen years ago, The Joint Commission mandated that health care organizations “encourage patients’ active involvement in their own care as a patient safety strategy,”^11^ and a linchpin of that requirement is data transparency and accessibility. Launched in 2010, the OpenNotes program, which allowed patients to access their electronic medical records, demonstrated that 99% of patients wanted the program to continue and 85% reported that access would inform their future provider and health system choices.^15^ In radiology, approaches such as leaving a summary statement at the end of the report,^16^ structured templates with standardized lexicon,^17,18^ and video reports^19^ have all been used to improve digital health literacy. Largely underexplored are emerging artificial intelligence (AI) tools to support patient understanding.

Using deep learning techniques, large language models (LLMs), such as OpenAI’s ChatGPT, Google Bard, and Microsoft Bing, have emerged as promising tools for the simplification of complex medical information.^20,21^ More specifically, these models leverage natural language processing (NLP) technologies to generate human-like text in response to a user’s prompts. To date, a comparative analysis of these LLMs in radiology has not been fully explored.

In this study, we compared the performance of several popular LLMs in producing simplified reports. Our objective was to evaluate the effectiveness of LLMs and provide insights into their potential for enhancing patient health literacy and promoting better patient–provider communication.

## Methods

To investigate the efficacy of four Large Language Models (LLMs) in simplifying radiology reports, we designed a comparative study focusing on OpenAI’s ChatGPT3.5 and ChatGPT4.0, Google Bard, and Microsoft Bing. Given that Bing has three conversational styles, we elected to use the precise setting over the creative or balanced settings. Our primary outcome was readability score, using an existing open-source dataset of reports.

### Dataset Selection and Modification

We used the MIMIC-III database, which is a comprehensive public database from the Beth Israel Deaconess Medical Center.^22,23^ A random selection of 254 anonymized reports was made to ensure representation of various examination types (MRI, CT, US (ultrasound), X-ray, Mammogram), anatomical regions, and lengths. This dataset allowed us to evaluate LLM performance across diverse clinical situations.

The reports in the datasets contained redacted information, so we altered the reports to state “Dr. Smith” where a physician name was redacted. Further, we changed redacted dates to “prior,” as many reports compared findings to previous studies.

### Prompt Selection

We first tested the prompt “Simplify this radiology report:” (Prompt 1). We then tested the prompt “I am a patient. Simplify this radiology report:” (Prompt 2).^24^ Lastly, we tested the prompt, “Simplify this radiology report at the 7^th^ grade level” (Prompt 3). Each prompt was followed with the radiology reports from the MIMIC-III database.

### Processing Radiology Reports and Readability Assessment

Each of the 254 radiology reports were processed individually by the 4 LLMs (accessed on May 1^st^, 2023: ChatGPT3.5 Legacy, ChatGPT4.0, Microsoft Bing, Google Bard) generating simplified versions of the original reports for each of the three prompts. In order to standardize the outputs and ensure equal comparison, we removed all formatting, including bullet points and numbered lists, as is consistent with previous readability studies.^25,26^ Ancillary information, such as “Sure I understand you would like a simplified version of your radiology report” and “please note I am not a medical professional,” was also removed to focus the analysis on the clinical content.

We assessed the LLMs’ ability to simplify complex radiology reports by employing four established readability indices: Gunning Fog (GF), Flesch-Kincaid Grade Level (FK), Automated Readability Index (ARI), and Coleman-Liau (CL) indices.^27^ Each index outputs a score which corresponds to a reading grade level (RGL). RGL relates directly to educational attainment: an RGL of 6 corresponds to a sixth-grade level, an RGL of 12 corresponds to a high school senior level, and an RGL of 17 corresponds to a four-year college graduate level.^28–31^

As previously described,^25^ we averaged the GF, FK, ARI, and CL readability scores for each output to calculate an averaged reading grade level score (aRGL). We applied the non-parametric Wilcoxon signed-rank and rank-sum tests to compare RGLs and aRGLs.

## Results

We tested the LLMs with the 3 distinct prompts across 5 imaging modalities: X-ray (N=45), US (N=11), MRI (N=47), CT (N=107), and mammogram (N=33). Original radiologist reports had a median aRGL of 17.2 overall, with X-rays at 13.7, ultrasounds at 14.6, MRIs at 16.5, CTs at 18.4, and mammograms at 18.8 (Table 1). When comparing original radiologist reports, X-ray reports were significantly more readable than CT, mammogram, and MRI reports (p<0.001), and ultrasound reports were significantly more readable than reports for CTs and mammograms (p<0.001, Suppl. Fig. 2). Despite these relative differences, original X-ray and ultrasound reports were still approximately at the college RGLs.

**Table 1:** Median of the aRGL for each LLM and prompt based on examination type.

| Model | Overall Median | CT Median | X-ray Median | MRI median | US median | Mammogram median |
| --- | --- | --- | --- | --- | --- | --- |
| Original Report | 17.2 | 18.4 | 13.7 | 16.5 | 14.6 | 18.8 |
| Prompt 1 \| ChatGPT3.5 | 10.5 | 10.8 | 10.4 | 10.2 | 10.8 | 9.8 |
| Prompt 2 ChatGPT3.5 | 7.6 | 7.8 | 8.5 | 7.2 | 6.5 | 7.0 |
| Prompt 3 ChatGPT3.5 | 6.7 | 6.9 | 8.0 | 6.6 | 4.6 | 5.5 |
| Prompt 1 ChatGPT4.0 | 10.5 | 10.1 | 10.6 | 11.7 | 8.3 | 10.8 |
| Prompt 2 ChatGPT4.0 | 8.0 | 7.9 | 8.7 | 8.0 | 8.6 | 7.0 |
| Prompt 3 ChatGPT4.0 | 7.0 | 7.2 | 9.1 | 6.7 | 6.3 | 5.5 |
| Prompt 1 Bard | 9.1 | 8.5 | 12.1 | 8.6 | 7.9 | 9.3 |
| Prompt 2 Bard | 12.3 | 12.4 | 10.6 | 13.4 | 14.3 | 12.9 |
| Prompt 3 Bard | 10.1 | 9.9 | 10.3 | 9.6 | 13.8 | 10.6 |
| Prompt 1 Bing | 9.4 | 8.1 | 12.6 | 11.4 | 6.6 | 9.6 |
| Prompt 2 Bing | 12.6 | 12.6 | 12.7 | 12.6 | 11.1 | 12.6 |
| Prompt 3 Bing | 11.6 | 11.2 | 11.8 | 12.0 | 11.4 | 11.8 |

All four LLMs significantly simplified original radiology reports from baseline complexity across all three prompts for MRI, CT, and mammogram (Figures 1-3, Suppl. Fig. 3). For X-ray and ultrasound, ChatGPT3.5, ChatGPT4.0, and Bing similarly achieved statistically significant simplification across all prompts, but Bard only simplified ultrasounds with Prompt 1 and X-rays with Prompt 2 and 3.

**Figure 1.**
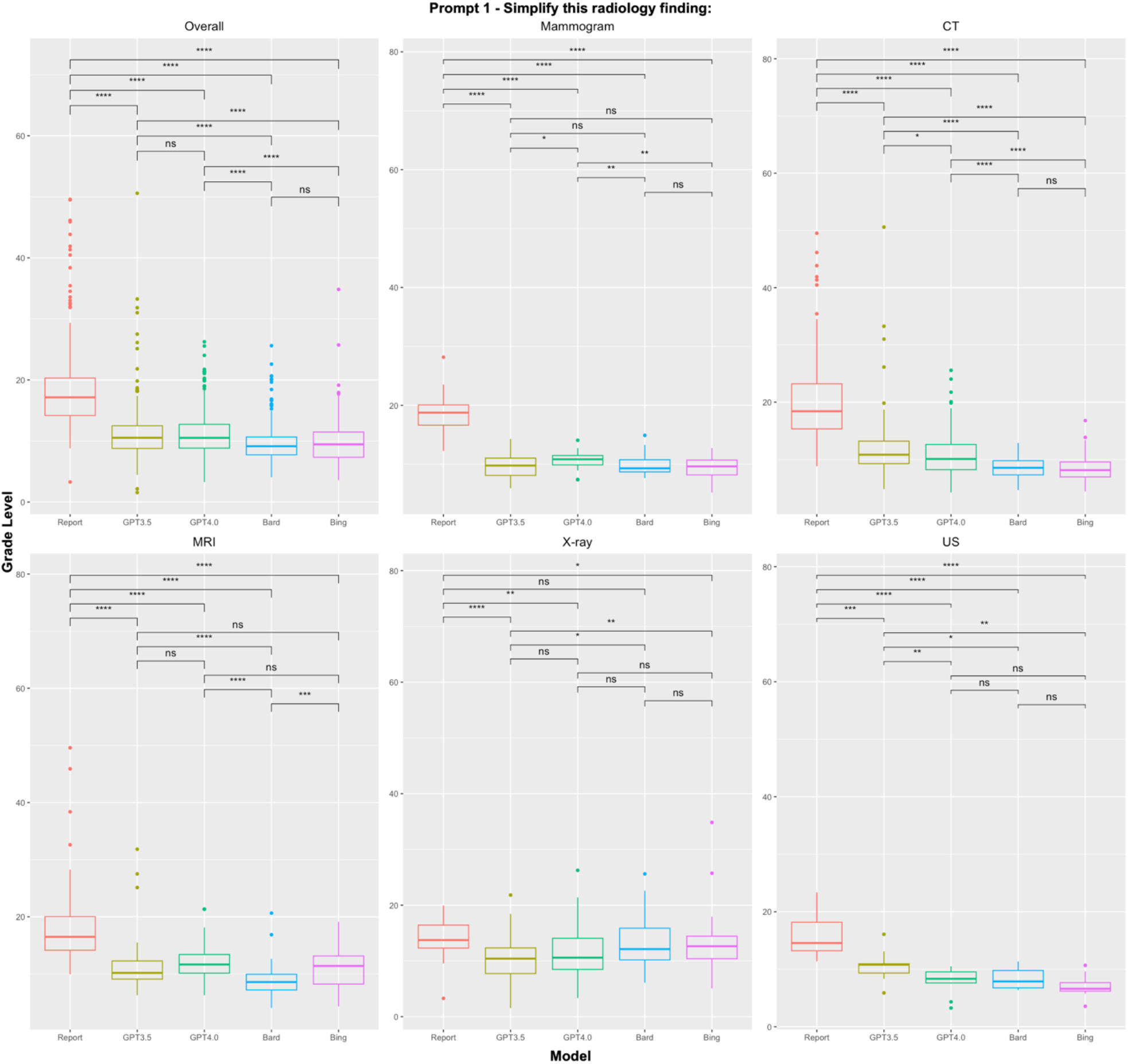
Readability scores of radiologist reports and LLMs using Prompt 1 – “Simplify this radiology finding:” *, **, ***, **** correspond to p<0.05, p<0.01, p<0.001, and p<0.0001, respectively.

**Figure 2.**
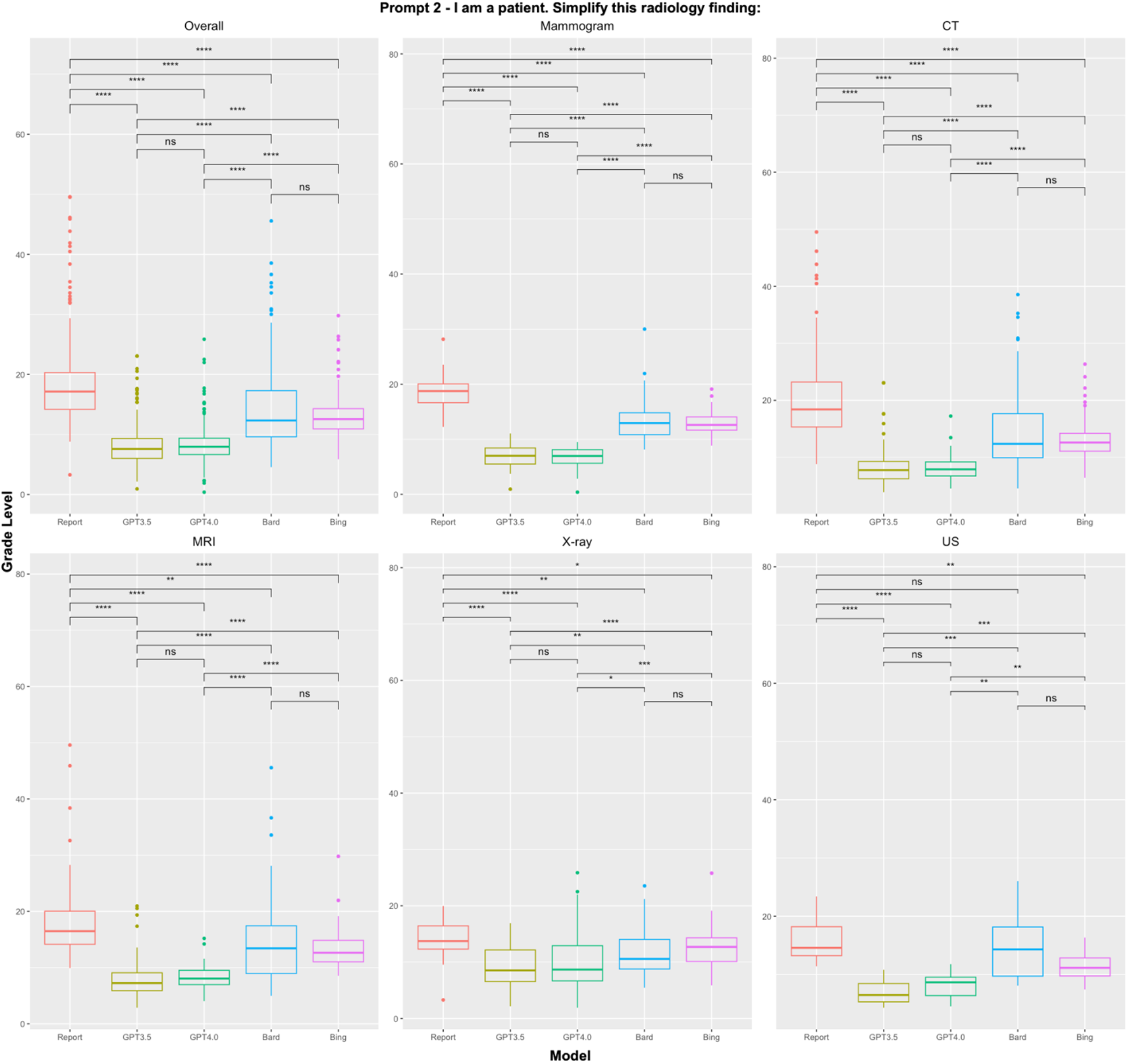
Readability scores of radiologist reports and LLMs using Prompt2 – “I am a patient. Simplify this radiology finding:” *, **, ***, **** correspond to p<0.05, p<0.01, p<0.001, and p<0.0001, respectively.

**Figure 3.**
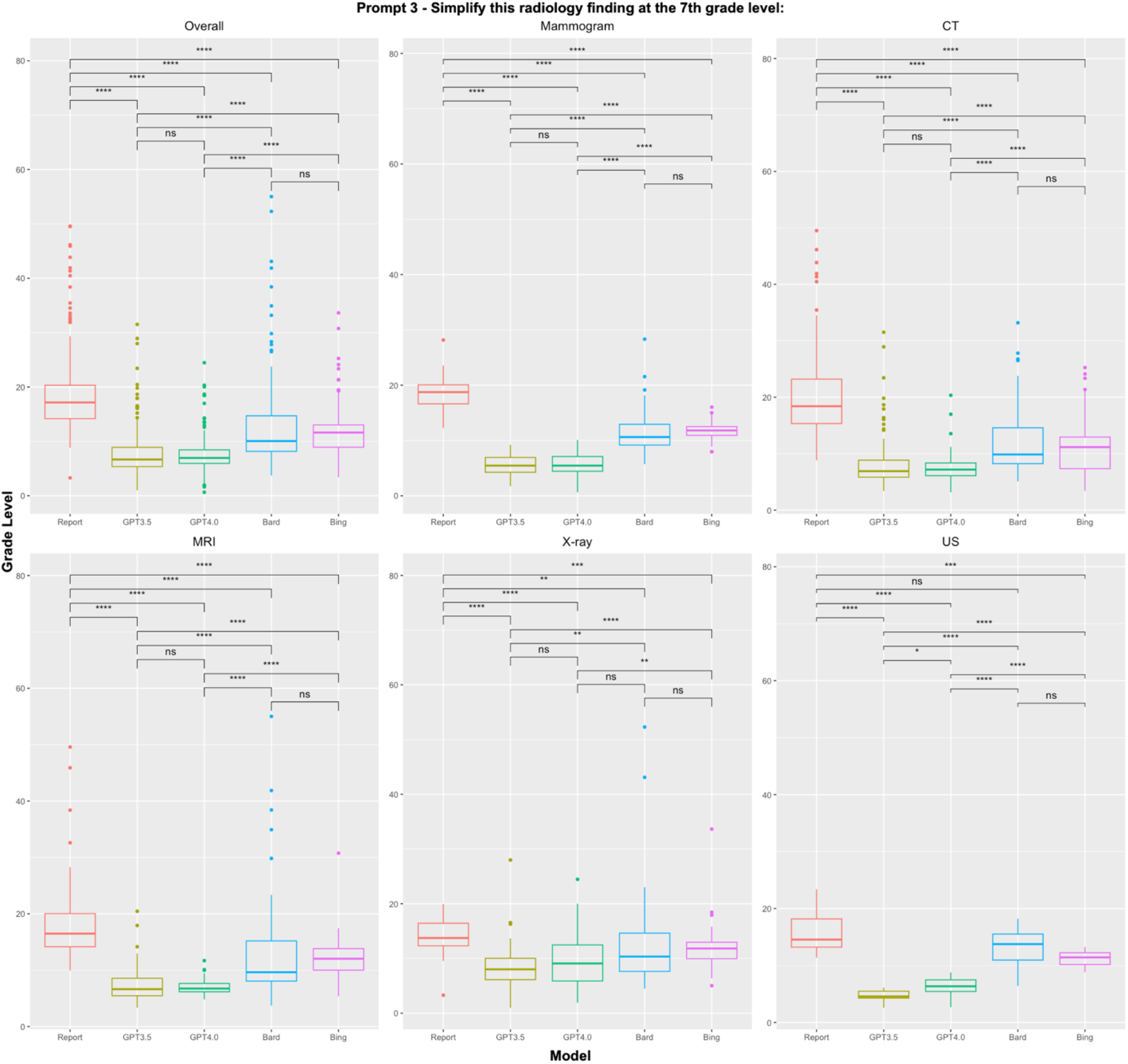
Readability scores of radiologist reports and LLMs using Prompt 3 – “Simplify this radiology finding at the 7th grade level:” *, **, ***, **** correspond to p<0.05, p<0.01, p<0.001, and p<0.0001, respectively.

### Prompt 1: “Simplify this radiology finding:”

Using Prompt 1, Bing and Bard achieved significantly lower combined median aRGL (9.4 and 8.1) than ChatGPT3.5 and ChatGPT4.0 (10.5 and 10.5, p<0.0001, Figure 1). Bard and Bing otherwise performed similarly, with Bard having the lowest combined median aRGLs for MRI (8.6, p<0.001), mammogram (9.3), and overall (9.1) reports and Bing for CT (8.1) and ultrasound (6.6). With Prompt 1, ChatGPT3.5 and ChatGPT4.0 performed similarly to each another, with typically higher aRGLs than Bing and Bard. The only exception was X-rays where ChatGPT3.5 had the lowest median aRGL (10.4), significantly lower than Bard and Bing.

### Prompt 2: “I am a patient. Simplify this radiology finding:”

With the added context of Prompt 2, ChatGPT3.5 and ChatGPT4.0 produced outputs with significantly lower aRGLs overall compared to Bard and Bing (p<0.0001) and for all imaging modalities tested (p<0.05, Figure 2). While there were no significant differences between ChatGPT3.5 and ChatGPT4.0, ChatGPT3.5 had the lowest median aRGL outputs for all imaging modalities (overall 7.6, CT 7.8, X-ray 8.5, MRI 7.2, US 6.5, and mammogram 7.0, Table 1).

### Prompt 3: “Simplify this radiology finding at the 7th grade level:”

Using Prompt 3 revealed similar outcomes to Prompt 2. The ChatGPT models significantly outperformed Bard and Bing overall and across all modalities (at least p<0.01, Figure 3), except for X-rays where no difference was found between Bard and ChatGPT4. Despite the two versions performing somewhat similarly, ChatGPT3.5 again produced the lowest aRGL outputs across our analysis (overall 6.7, CT 6.9, X-ray 8.0, MRI 6.6, ultrasound 4.6, and mammogram 5.5; Table 1).

### Prompt 1 vs Prompt 2 vs Prompt 3

Finally, we analyzed performance for each LLM across the three prompt combinations (Fig. 4). The ChatGPT models performed better at reducing aRGL with Prompt 2 and Prompt 3 than with Prompt 1 (p<0.0001); Prompt 3 also outperformed Prompt 2 (p<0.01). On the other hand, Bard and Bing performed better with Prompt 1 when compared to Prompt 2 and Prompt 3 (p<0.0001). We also observed that Prompt 3 outperforms Prompt 2 in producing lower aRGL outputs for Bard and Bing as well (p<0.0001).

**Figure 4.**
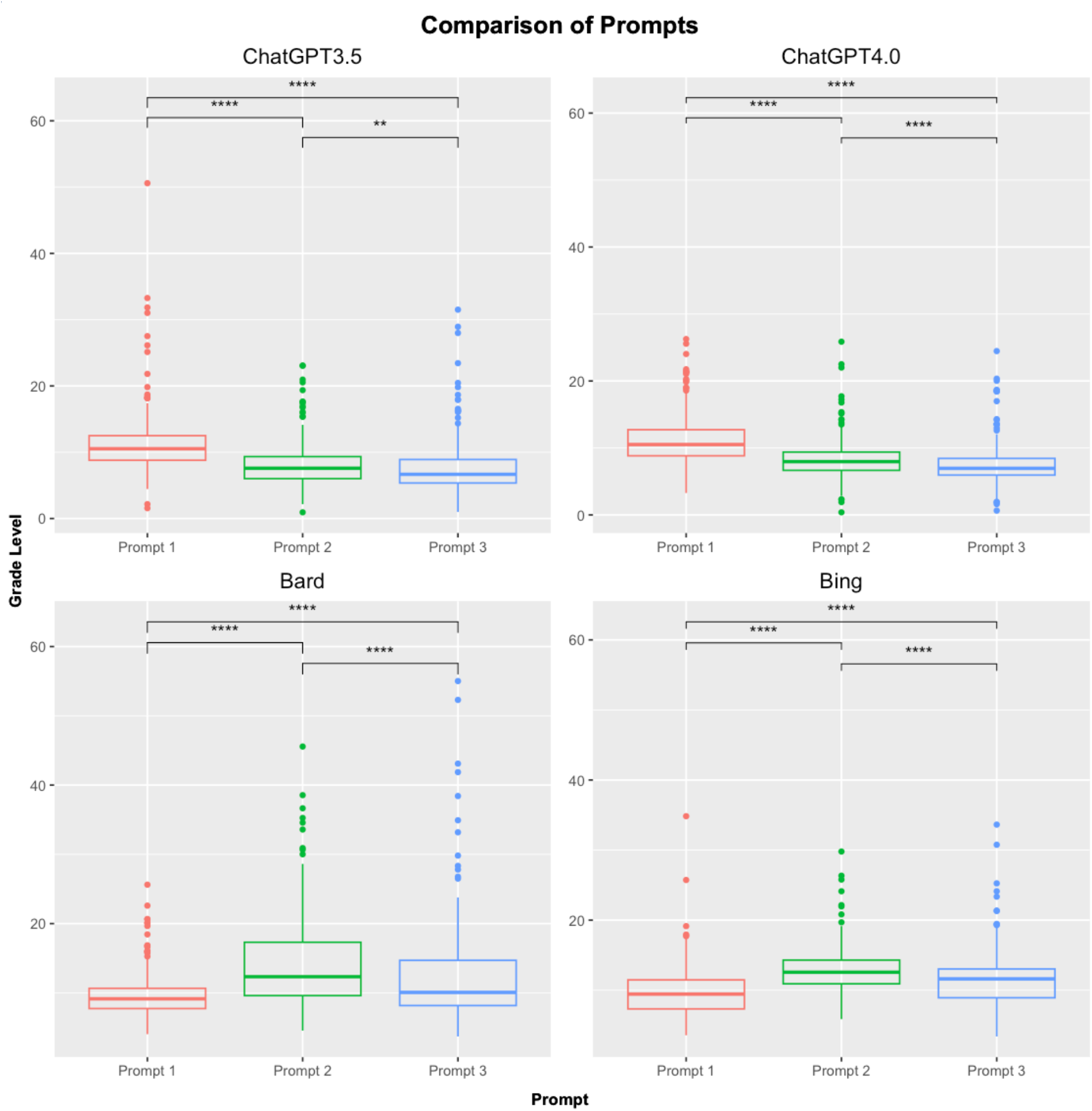
Comparison of each prompt within LLM. *, **, ***, **** correspond to p<0.05, p<0.01, p<0.001, and p<0.0001, respectively.

## Discussion

In this study, we showed that the baseline readability of radiology reports across CT, X-ray, MRI, ultrasound, and mammograms are above the college graduate level but OpenAI’s ChatGPT3.5 and ChatGPT4.0, Google Bard, and Microsoft Bing can all successfully simplify these reports. The success of each of the LLMs varied, however, according to the specific prompt wording. Microsoft Bing and Google Bard performed best with a straightforward request to simplify a radiology report (Prompt 1), while the ChatGPT models performed best when provided with added context, such as the user specifying they were a patient (Prompt 2) or requesting simplification at the 7^th^ grade level (Prompt 3).

Out of countless potential prompts that could have been tested, we focused our analysis on these three to determine how different types of context impacted readability. Prompt 1 was the simplest, specifying only that the inputted text will be a radiology report and that the LLM is tasked with simplifying it. The other two prompts offered additional context. For Prompt 3, we specified the 7^th^ grade level because the American Medical Association and National Institutes of Health recommend that patient education materials should be written between the third- and seventh-grade levels given that the average American reads at the eighth-grade level.^18,32,33^ As expected, Prompt 3 outperformed Prompt 2 across all LLMs tested, although we recognize that requesting simplification at a specific grade level is less accessible for most users than specifying that “I am a patient.” Unexpectedly, however, Prompt 1 obtained the lowest aRGLs for two of the four LLMs tested, Microsoft Bing and Google Bard,—suggesting that richer context does not always equate to improved readability for every LLM.

Several explanations may underlie the observed differences in readability scores across the LLMs. For one, variations in training data and preprocessing techniques could impact the different LLMs’ ability to handle the jargon, abbreviations, and numerical information found in radiology reports.^34^ Furthermore, there may simply be fundamental differences in LLM architectures and algorithms that make certain models more amenable to simplifying medical information.^35^ We nonetheless found the differences between Microsoft Bing and Open AI’s ChatGPT models remarkable because Bing is powered by OpenAI. The finding that ChatGPT3.5 produced similar outputs to ChatGPT4.0 was also notable because it suggests that updated software does not automatically equate to improved performance, at least in regards to readability.

With patients already using these LLMs to simplify medical information,^36^ providers cannot ignore how the information-sharing landscape has changed and should consider accordingly. For instance, radiologists may consider using LLMs proactively to create a patient-friendly report, inputting it into the electronic medical record alongside their original report to help alleviate patient anxiety, misunderstanding, and emotional distress.^37^ Epic, Cerner, and other electronic health record companies may soon integrate LLMs into their software such that radiologists would not need to leave the interface to rely on third party tools.^38^

While LLMs demonstrate promise in helping patients better understand their radiology reports, the ultimate goal should be to strike a balance between readability and preserving clinical fidelity.^39^ Indeed, excessive simplification could contribute to clinical inaccuracies and actually cause patients greater anxiety, so the role of healthcare providers in facilitating communication and understanding should not be overlooked. We believe LLMs could eventually be used as supplementary tools to aid patient-provider communication rather than a replacement for personal interaction and discussion, however, it is essential to study the accuracy and fidelity of these outputs before recommending their usage on a wider-scale.^40^

This study has limitations. For one, radiologists or medical professionals did not assess simplified outputs, so we cannot speak to the accuracy, fidelity, and clinical utility of these reports. The readability metrics used in this study are similarly limited because they are language- and structure-focused, so these measures do not necessarily capture relevance or comprehensibility from a medical perspective. Furthermore, due to the formulaic nature of these metrics, outputted RGLs were sometimes above a meaningful grade level (i.e., a score of 30) and thus held little interpretability on their own. In this study, we were interested in assessing the readability of reports after LLM simplification and evaluating relative differences from baseline. Finally, we extracted radiology reports from the MIMIC-III dataset, which is derived from a single hospital, and employed a cross-sectional design, which may not be ideal for capturing continuous changes in LLMs’ performance. A longitudinal study design, as well as a larger, more diverse dataset, might have improved these results’ validity and generalizability.

## Conclusion

Our study highlights how radiology reports are complex medical documents that implement language and style above the college graduate reading level, but LLMs are powerful tools for simplifying these reports. Our findings should not be viewed as an endorsement for any particular LLM, instead demonstrating that each LLM tested has the ability to simplify radiology reports across modalities. Careful fine-tuning and customization for each LLM may ensure optimal simplification while maintaining the clinical integrity of the reports.

## Data Availability

All data produced in the present study are available upon reasonable request to the authors.

---

**eFigure 1:**
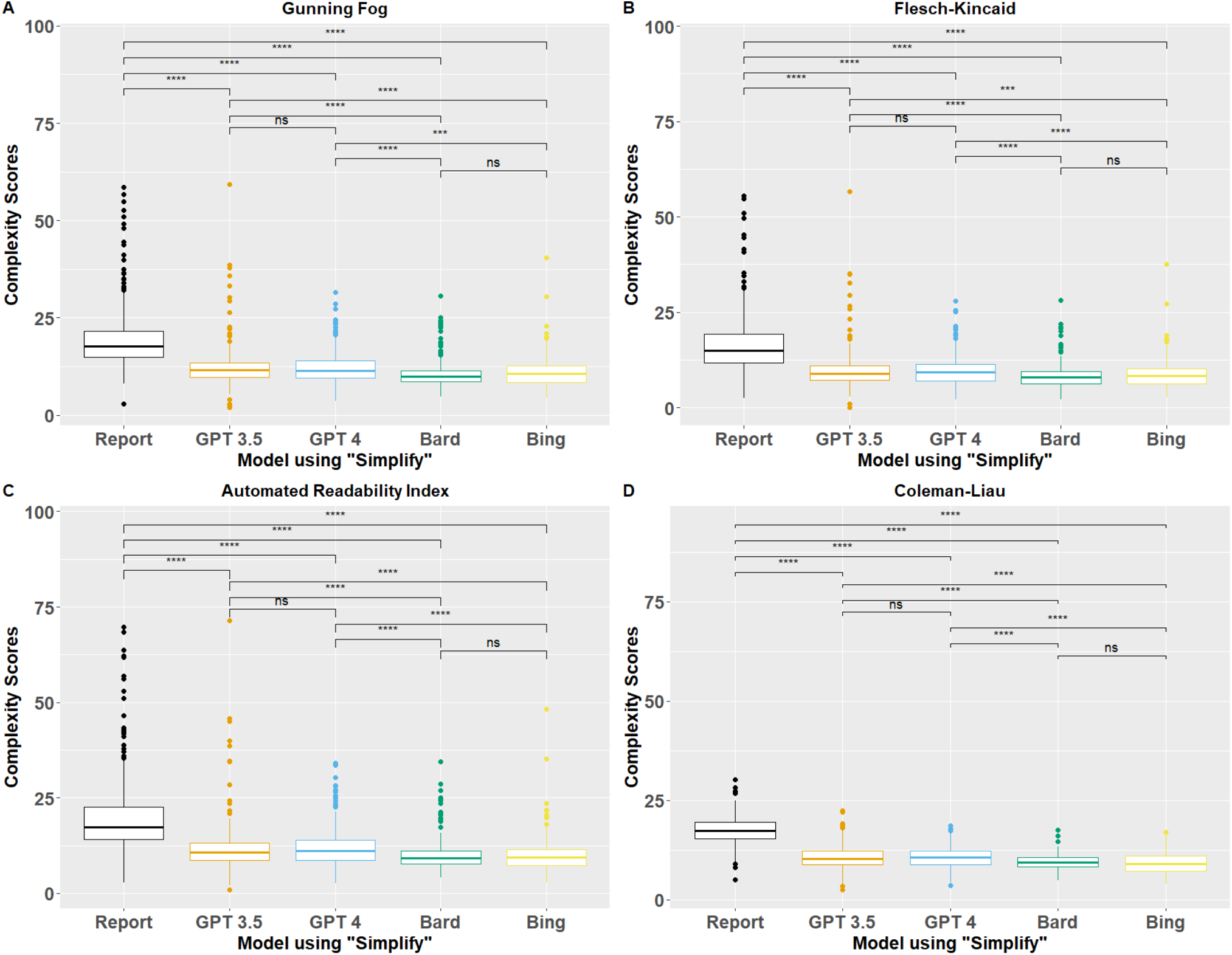
GF, FK, ARI, and CL readability scores using Prompt 1. *, **, ***, **** correspond to p<0.05, p<0.01, p<0.001, and p<0.0001, respectively

**eFigure 2:**
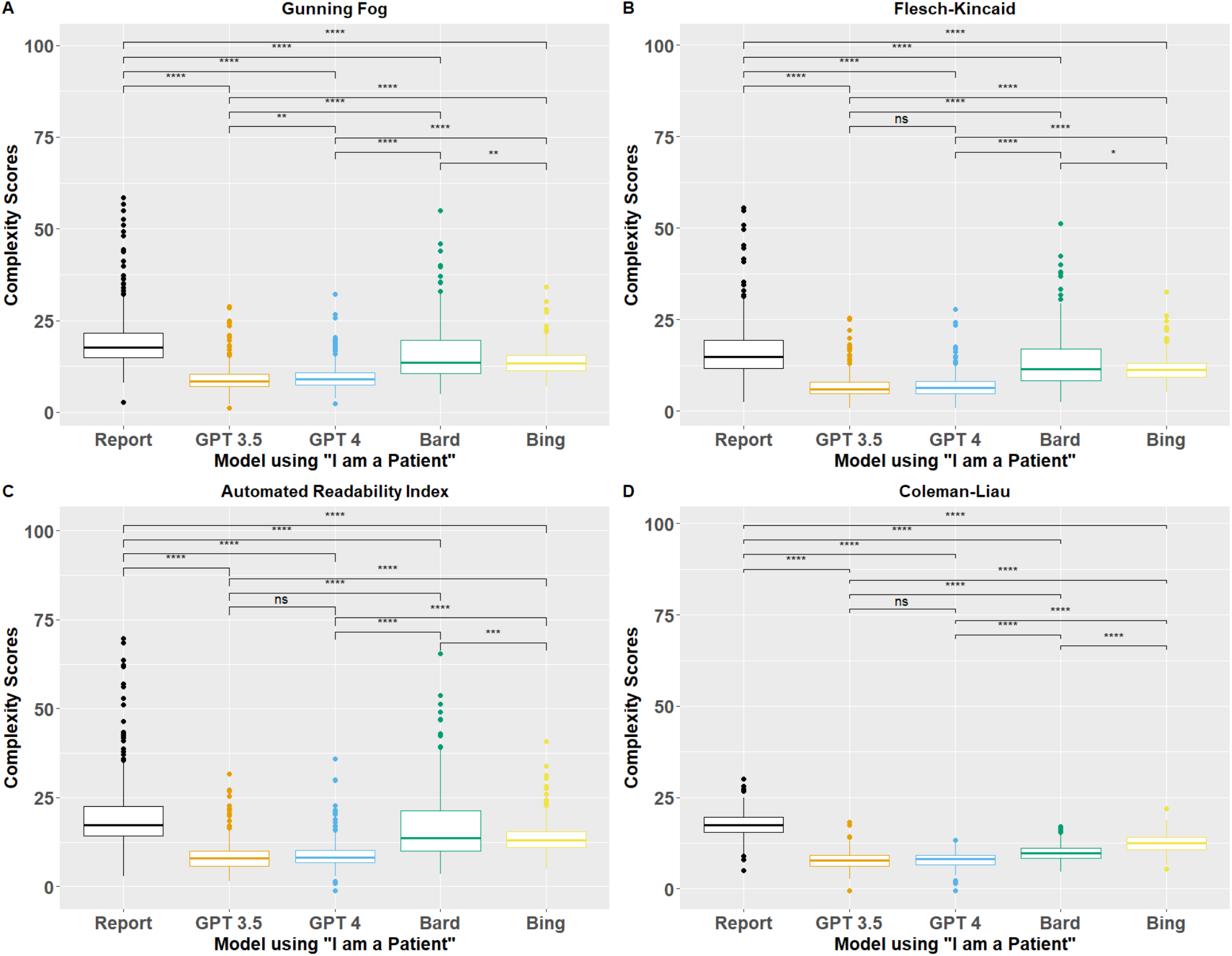
GF, FK, ARI, and CL readability scores using Prompt 2. *, **, ***, **** correspond to p<0.05, p<0.01, p<0.001, and p<0.0001, respectively

**eFigure 3:**
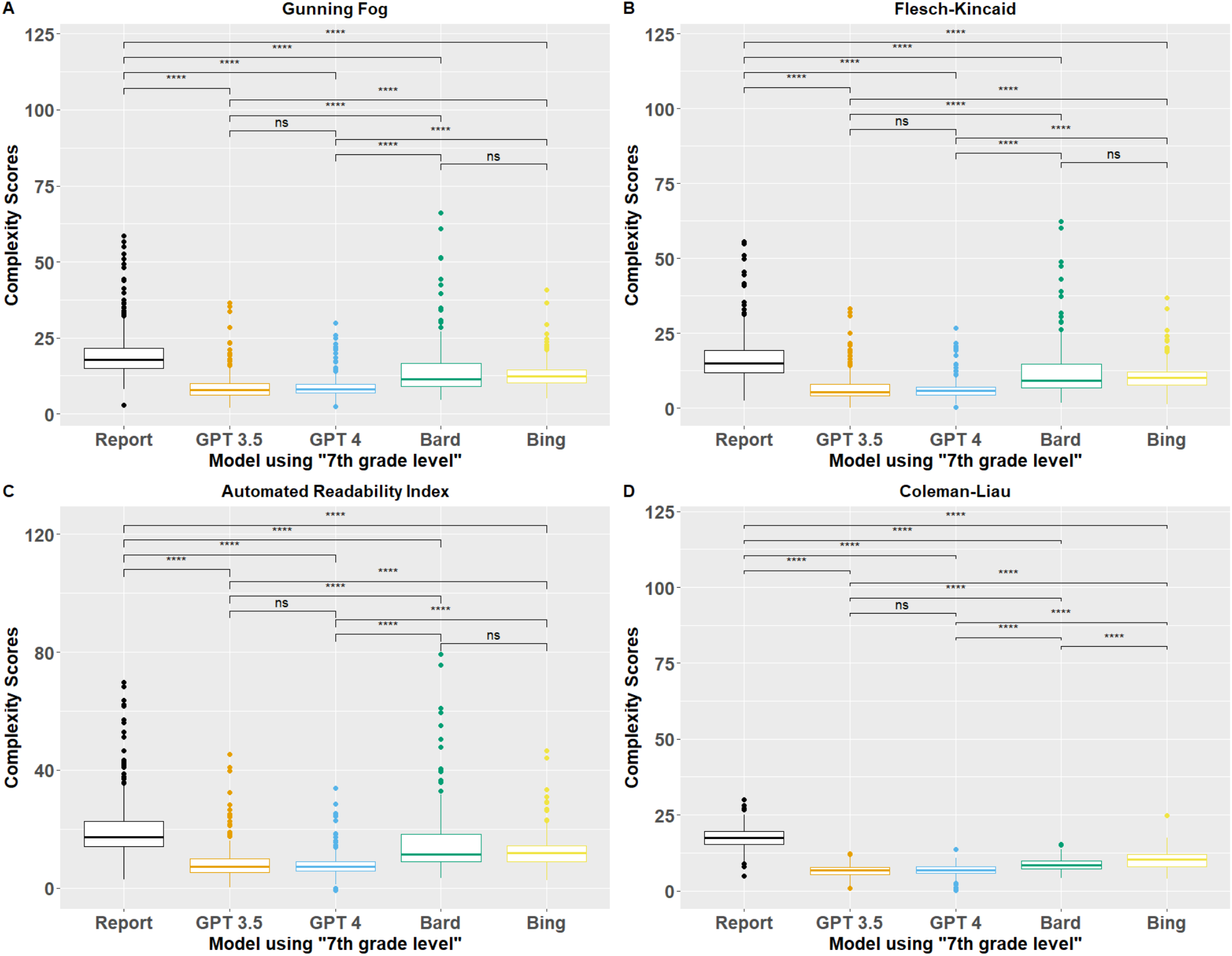
GF, FK, ARI, and CL readability scores using Prompt 3. *, **, ***, **** correspond to p<0.05, p<0.01, p<0.001, and p<0.0001, respectively

**eTable 1:**
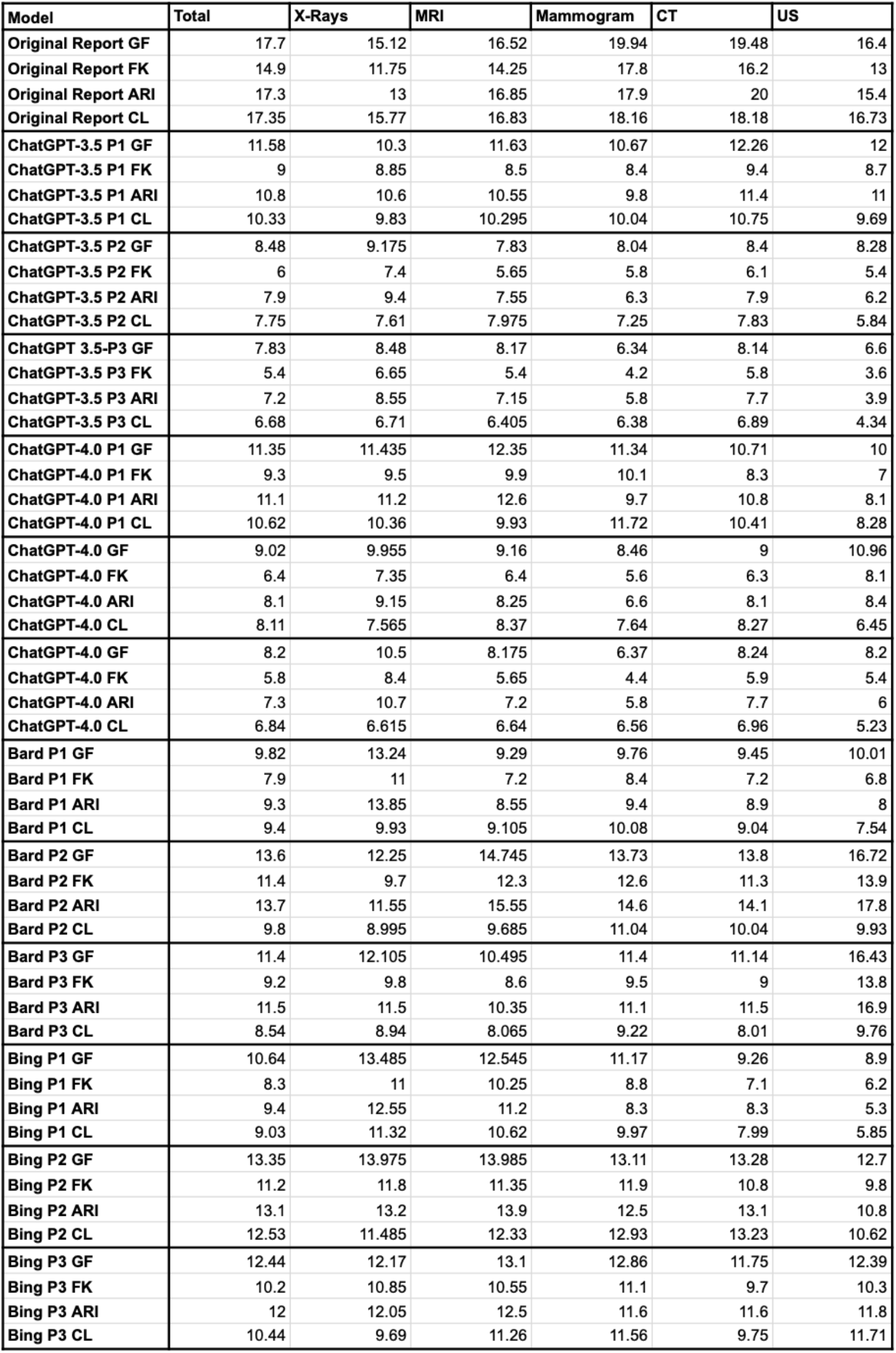
Median scores across LLM, prompt, modality, and readability index. *, **, ***, **** correspond to p<0.05, p<0.01, p<0.001, and p<0.0001, respectively

**eTable 2:**
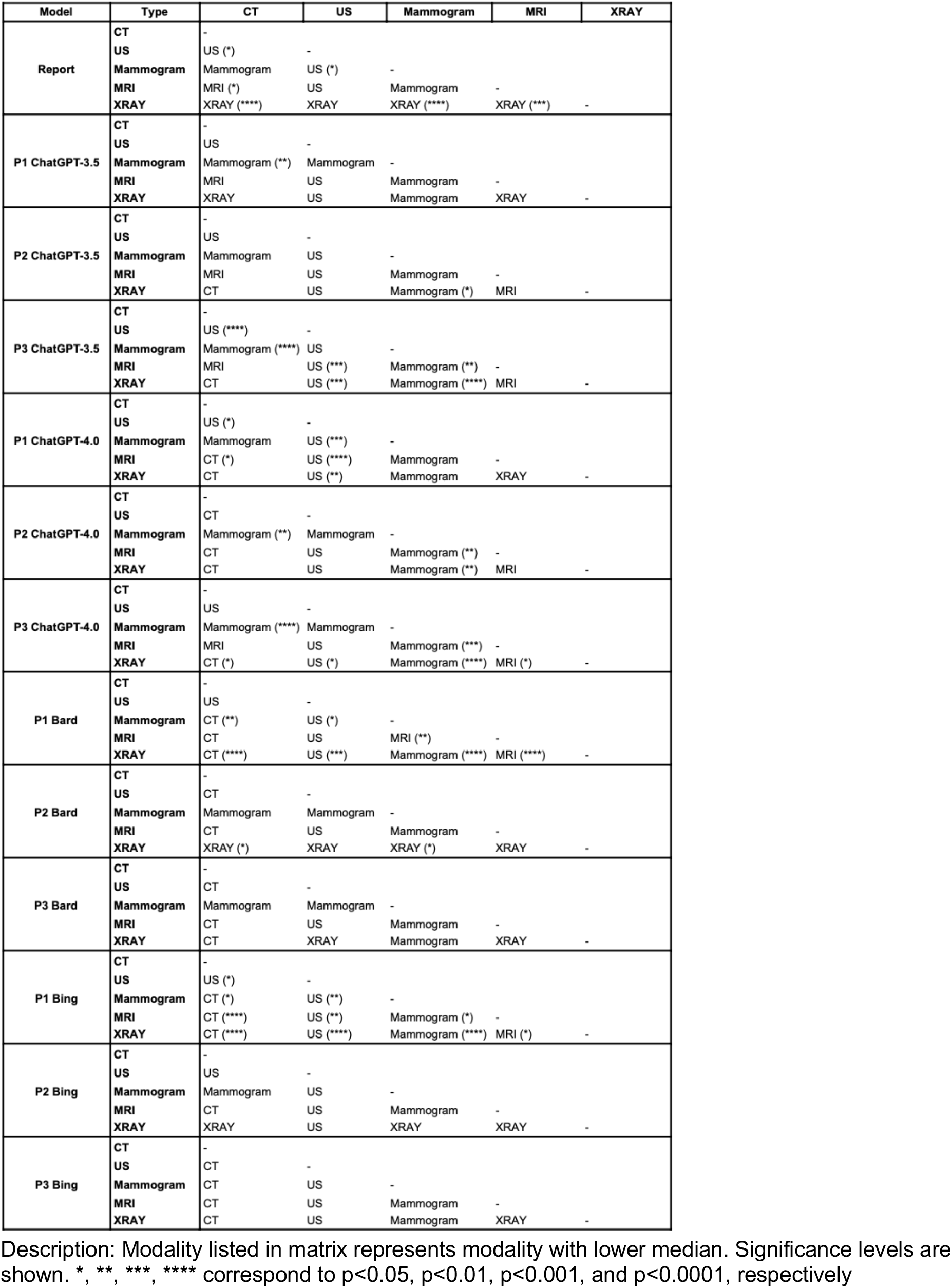
Comparison of each modality within each prompt and LLM.

**eTable 3:**
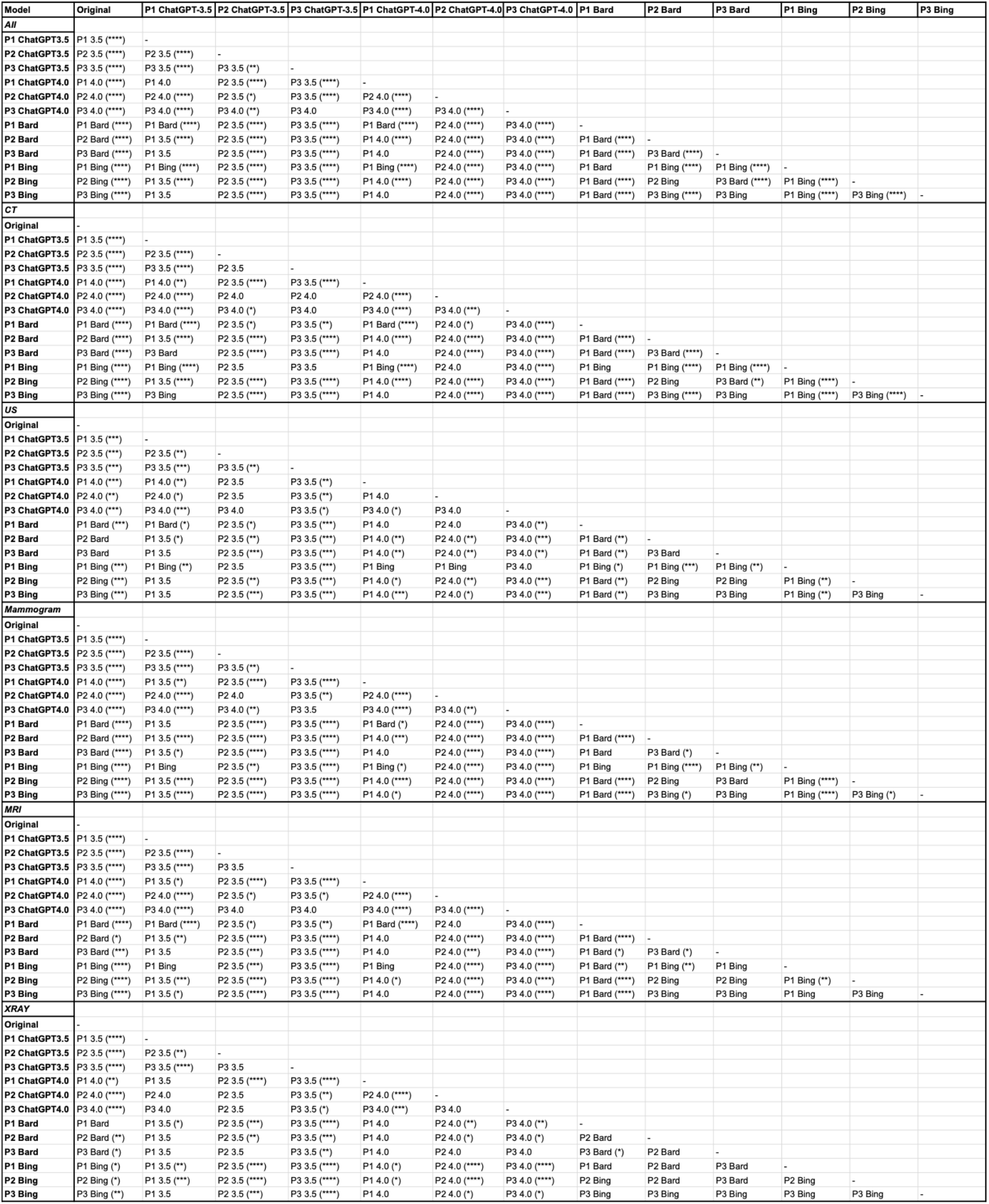
Comparison of each prompt and LLM combination within modality. Description: P1-Prompt 1, P2 – Prompt 2, P3 – Prompt 3. Combination listed in matrix has lower median; significance levels are shown. *, **, ***, **** correspond to p<0.05, p<0.01, p<0.001, and p<0.0001, respectively

